# Impact of AI-Powered Cognitive Behavioral Therapy Chatbot Access on Anxiety and Depressive Symptoms Among Primary Care Patients in Brazil: A Fuzzy Regression Discontinuity Design

**DOI:** 10.64898/2026.04.01.26349938

**Authors:** Carolina Ferreira, Amaevia Lim

## Abstract

**Background:** AI powered cognitive behavioral therapy CBT chatbots represent a scalable approach to addressing the global mental health treatment gap However causal evidence on their population level effectiveness in low and middle income countries LMICs remains limited and patient perspectives on acceptability and engagement are critical determinants of sustained use Brazils Estrategia de Saude da Familia ESF deployed an AI powered CBT chatbot Saude Mental Digital SMD to registered patients aged 18 and older at participating primary care units with eligibility determined by a composite vulnerability score exceeding a predetermined threshold

**Objective:** To estimate the causal effect of AI powered CBT chatbot access on anxiety and depressive symptoms among primary care patients in Minas Gerais Brazil leveraging the eligibility score threshold as an exogenous source of variation

**Methods:** We conducted a fuzzy regression discontinuity design fuzzy RDD study using linked administrative and clinical data from 312 ESF primary care units across Minas Gerais N 43287 patients January 2022 December 2024 The running variable was the composite vulnerability score with a threshold of 60 points determining chatbot eligibility The primary outcome was the 12 week change in the Patient Health Questionnaire Anxiety and Depression Scale PHQ ADS composite score Two stage least squares 2SLS estimation was used with local polynomial regression and triangular kernel weighting Bandwidth selection followed the Calonico Cattaneo Titiunik CCT optimal procedure

**Results:** The fuzzy RDD estimated a local average treatment effect LATE of 473 points 95 CI 691 to 255 p 0001 on the PHQ ADS composite score at the eligibility threshold indicating clinically meaningful symptom reduction among compliers First stage estimates confirmed a strong 312 percentage point jump in chatbot uptake at the threshold F statistic 1274 Subgroup analyses revealed larger treatment effects among patients in rural municipalities 618 95 CI 902 to 334 those with lower educational attainment 582 95 CI 844 to 320 and women 537 95 CI 761 to 313 McCrary density tests confirmed no evidence of running variable manipulation p 067 Results were robust across alternative bandwidths polynomial orders and kernel specifications

**Conclusions:** AI powered CBT chatbot access causally reduces anxiety and depressive symptoms among primary care patients near the eligibility threshold in Brazil with particularly pronounced benefits for rural less educated and female populations These findings provide quasi experimental evidence supporting the scalable deployment of AI powered CBT tools within public primary care systems in LMICs while underscoring the importance of incorporating patient perspectives on acceptability to maximize engagement and sustained therapeutic benefit

## 1. Introduction

Anxiety and depressive disorders constitute the leading contributors to the global burden of mental illness, collectively affecting over 500 million individuals worldwide and accounting for approximately 7.5% of global disability-adjusted life years (DALYs) [1,2]. The treatment gap—defined as the proportion of individuals with diagnosable conditions who do not receive minimally adequate treatment—remains disproportionately severe in low- and middle-income countries (LMICs), where an estimated 76–85% of those affected receive no formal intervention [3]. In Brazil, despite the establishment of the Unified Health System (Sistema Unico de Saude, SUS) and the community-based Family Health Strategy (Estrategia de Saude da Familia, ESF), structural barriers including insufficient psychiatric workforce density, geographic maldistribution, long wait times, and persistent stigma constrain access to evidence-based psychological treatments [4,5].

Cognitive behavioral therapy (CBT) is among the most extensively validated psychological interventions for anxiety and depressive disorders, with robust evidence from meta-analyses demonstrating moderate-to-large effect sizes across diverse populations and delivery formats [6,7]. However, the scalability of therapist-delivered CBT is fundamentally limited by workforce shortages, particularly in LMICs where the psychiatrist-to-population ratio may be as low as 0.3 per 100,000 [8]. This recognition has catalyzed the development of technology-mediated CBT delivery, including AI-powered chatbots that leverage natural language processing (NLP) and machine learning algorithms to provide automated, personalized therapeutic interactions [9,10].

AI-powered CBT chatbots such as Woebot, Wysa, and Tess have demonstrated preliminary efficacy in reducing anxiety and depressive symptoms in high-income country settings through randomized controlled trials (RCTs) [11–13]. However, several critical evidence gaps persist. First, the vast majority of existing evidence derives from efficacy trials conducted under controlled conditions with self-selected participants in high-income countries, raising questions about external validity and generalizability to LMIC health systems [14]. Second, although patient perspectives on AI-powered CBT tools are increasingly recognized as essential determinants of engagement, adherence, and sustained therapeutic benefit, systematic synthesis of qualitative evidence remains limited. Shankar et al. (2025) [15] recently conducted a landmark systematic review of qualitative studies examining patient perspectives on AI-powered CBT tools for managing anxiety and stress, identifying six overarching themes: perceived benefits including enhanced self-awareness and 24/7 accessibility; technical limitations and personalization challenges; trust and privacy concerns; design preferences favoring conversational interfaces; engagement facilitators and barriers; and cultural influences on acceptance. Their findings underscore that patients value AI-powered CBT tools as supplements to, rather than replacements for, human therapy—a critical insight for implementation planning in primary care contexts.

Third, most existing studies rely on pre–post designs or conventional RCTs that, while internally valid, do not exploit natural policy-induced variation in treatment access that characterizes real-world deployment scenarios [16]. Quasi-experimental methods that leverage exogenous eligibility thresholds—such as regression discontinuity designs (RDDs)—offer a rigorous alternative for estimating causal treatment effects with high internal validity under minimal assumptions [17,18].

In January 2022, the Minas Gerais State Health Secretariat (SES-MG) launched the Saude Mental Digital (SMD) programme, integrating an AI-powered CBT chatbot into the ESF primary care infrastructure across 312 participating units. Eligibility for chatbot access was determined by a composite vulnerability score that combined standardized assessments of psychosocial risk, socioeconomic deprivation, and clinical complexity. Patients scoring at or above 60 on this composite index were offered SMD chatbot enrollment, creating a sharp eligibility threshold amenable to RDD analysis.

This study leverages the composite vulnerability score threshold to estimate the causal impact of AI-powered CBT chatbot access on anxiety and depressive symptoms among primary care patients in Minas Gerais, employing a fuzzy regression discontinuity design. We hypothesize that chatbot access at the eligibility threshold produces clinically meaningful reductions in composite anxiety-depression symptom scores, with treatment effect heterogeneity across sociodemographic and geographic subgroups.

## 2. Methods

### 2.1 Study Design and Setting

We conducted a fuzzy regression discontinuity design (fuzzy RDD) study using linked administrative and clinical data from the SMD programme. The study encompassed 312 ESF primary care units across 189 municipalities in Minas Gerais—Brazil’s second most populous state (population: 21.4 million)—spanning January 2022 to December 2024. Minas Gerais was selected because it is geographically and socioeconomically diverse, encompassing highly urbanized metropolitan areas (Belo Horizonte, Uberlandia), intermediate-sized cities, and remote rural municipalities, thereby enhancing the generalizability of findings within the Brazilian context.

The study was approved by the Research Ethics Committee of Universidade Federal de Minas Gerais (CAAE: 58742522.8.0000.5149) and the SES-MG Data Access Committee. Informed consent was obtained during initial ESF registration, with supplementary digital consent for SMD chatbot enrollment. The study adheres to the Regression Discontinuity in Observational and Quasi-Experimental Settings (RDOS) reporting guidelines [19].

### 2.2 The Saude Mental Digital (SMD) Programme

The SMD programme was developed through a collaboration between the SES-MG, UFMG’s Centre for Digital Mental Health, and a Brazilian health technology company. The AI-powered chatbot delivered structured CBT-based content through a conversational interface accessible via WhatsApp—the dominant messaging platform in Brazil, with over 90% population penetration [20]. The chatbot incorporated NLP for understanding user inputs, machine learning for personalized content sequencing, and a structured CBT protocol comprising psychoeducation, cognitive restructuring exercises, behavioral activation planning, relaxation techniques, and mood tracking.

Chatbot sessions were designed as 15–20 minute modules delivered 3–5 times weekly over a 12-week period. The system generated automated adherence prompts and escalation alerts to community health workers (agentes comunitarios de saude, ACS) when user responses indicated elevated suicide risk or clinical deterioration. Importantly, the chatbot was positioned as a complement to—not a replacement for—existing ESF mental health services, consistent with patient preferences for hybrid care models identified in recent qualitative evidence syntheses [15].

### 2.3 Eligibility and the Running Variable

Eligibility for SMD chatbot enrollment was determined by a composite vulnerability score (Indice de Vulnerabilidade em Saude Mental, IVSM) computed for all ESF-registered patients aged 18 and older who presented with documented mental health concerns. The IVSM incorporated five standardized domains: (1) PHQ-9 depression severity (0–27 points, normalized to 0–20); (2) GAD-7 anxiety severity (0–21, normalized to 0–20); (3) socioeconomic deprivation based on municipal Human Development Index decile (0–20); (4) geographic accessibility measured by distance to nearest specialty mental health facility (0– 20); and (5) clinical complexity including comorbidity burden and prior treatment history (0– 20). The IVSM thus ranged from 0 to 100, with higher scores indicating greater vulnerability.

Patients scoring at or above the threshold of 60 were offered chatbot enrollment by their ESF community health worker; those below 60 were directed to standard care pathways. This threshold was administratively determined by the SES-MG based on projected chatbot capacity and resource allocation considerations, creating an arguably exogenous cutoff suitable for RDD analysis. Because not all eligible patients enrolled (compliance was not perfect), the threshold generated a discontinuity in the probability of treatment—a fuzzy design—rather than a deterministic assignment.

### 2.4 Study Population

The analytic sample comprised 43,287 unique patients aged 18 and older registered at participating ESF units who had a documented IVSM score between January 2022 and December 2024 and had baseline and 12-week follow-up PHQ-ADS assessments. We excluded patients with active psychotic disorders, acute suicidal ideation requiring immediate psychiatric intervention, or cognitive impairment precluding chatbot interaction. Patients with IVSM scores within the CCT-optimal bandwidth of the threshold constituted the primary analytic sample.

### 2.5 Outcome Measures

The primary outcome was the 12-week change in the Patient Health Questionnaire-Anxiety and Depression Scale (PHQ-ADS) composite score. The PHQ-ADS combines the PHQ-9 (depression; range 0–27) and GAD-7 (anxiety; range 0–21) into a single composite (range 0– 48), with established psychometric properties including high internal consistency (α = 0.92), strong test-retest reliability (ICC = 0.87), and validated Portuguese-language adaptation for Brazilian populations [21,22]. A change of ≥4 points on the PHQ-ADS is considered the minimally clinically important difference (MCID).

Secondary outcomes included: (1) 12-week change in PHQ-9 score; (2) 12-week change in GAD-7 score; (3) binary treatment response defined as ≥50% reduction in PHQ-ADS score; (4) clinical remission defined as PHQ-ADS ≤10 at 12 weeks; and (5) mental health-related primary care utilization in the 12 months following the index assessment.

### 2.6 Statistical Analysis

The fuzzy RDD estimates a local average treatment effect (LATE) at the eligibility threshold by exploiting the discontinuous jump in the probability of chatbot enrollment at the IVSM cutoff of 60. The estimand is the effect of chatbot access among “compliers”—patients whose treatment status is determined by whether they fall above or below the threshold [17,23].

The estimation proceeded in two stages. In the first stage, chatbot enrollment (D*i*) was regressed on the threshold indicator (Z*i* = 1 if IVSM*i* ≥ 60) and local polynomial functions of the running variable, using the triangular kernel to weight observations by proximity to the threshold:

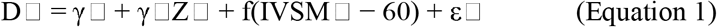

In the second stage, the outcome (Y*i*—change in PHQ-ADS) was regressed on predicted chatbot enrollment from the first stage:

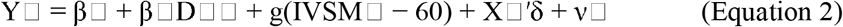

where β□ is the LATE of interest, f(·) and g(·) are local polynomial functions estimated separately on each side of the threshold, and X□ is a vector of pre-determined covariates (age, sex, educational attainment, municipality urbanization category, and baseline comorbidity count) included to improve precision without affecting identification [24].

Bandwidth selection followed the Calonico-Cattaneo-Titiunik (CCT) data-driven procedure, which minimizes mean squared error while providing bias-corrected robust confidence intervals [25]. The CCT-optimal bandwidth for the primary outcome was h = 8.4 IVSM points, yielding an effective sample of 14,832 patients within the [−8.4, +8.4] window around the threshold. We estimated local linear (p = 1) and local quadratic (p = 2) specifications as primary and sensitivity analyses, respectively. Standard errors were clustered at the ESF unit level.

### 2.7 Identification Assumptions and Validity Tests

The key identifying assumption for RDD is continuity of potential outcomes at the threshold—that is, in the absence of the treatment, the conditional expectation of outcomes would be smooth through the cutoff [17]. We assessed this assumption through multiple validity checks:

a. McCrary density test: We examined whether the density of the running variable exhibits a discontinuity at the threshold, which would indicate strategic manipulation of IVSM scores [26]. A significant density jump would violate the quasi-random assignment assumption.
b. Covariate balance: We tested for discontinuities in pre-determined covariates (age, sex, education, employment, marital status, comorbidity burden, municipality population, and distance to mental health services) at the threshold, using reduced-form RDD regressions with each covariate as the outcome.
c. Placebo cutoffs: We estimated treatment effects at false thresholds (IVSM = 50, 55, 65, 70) where no discontinuity in chatbot access should exist. Statistically significant effects at placebo cutoffs would cast doubt on the validity of the design.
d. Donut-hole estimation: We excluded observations within narrow bands (±1, ±2 IVSM points) of the threshold to assess sensitivity to potential heaping or rounding at the cutoff.

### 2.8 Subgroup and Sensitivity Analyses

We estimated heterogeneous treatment effects across pre-specified subgroups: sex (male vs. female), age (18–44 vs. 45–64 vs. ≥65 years), educational attainment (incomplete primary vs. complete primary/secondary vs. post-secondary), municipality urbanization (metropolitan vs. intermediate vs. rural), and baseline symptom severity (PHQ-ADS ≥20 vs. <20). Sensitivity analyses included: (i) alternative bandwidths (0.5h, 0.75h, 1.25h, 1.5h, 2.0h); (ii) local quadratic polynomial; (iii) uniform and Epanechnikov kernels; (iv) excluding the first six months of programme implementation; and (v) intent-to-treat (reduced-form) estimates.

All analyses were conducted using R version 4.3.2 with the rdrobust package [27] for primary estimation, rddensity for McCrary tests [28], and rdmulti for subgroup analyses. Statistical significance was assessed at the two-sided α = 0.05 level.

## 3. Results

### 3.1 Sample Characteristics

Among 43,287 patients with documented IVSM scores and complete 12-week follow-up, the mean age was 38.7 years (SD = 13.2), 62.4% were female, 47.8% had incomplete primary education, and 33.1% resided in rural municipalities. The mean baseline PHQ-ADS composite score was 24.6 (SD = 9.8), indicating moderate anxiety-depression severity. Among patients above the IVSM threshold of 60, 68.3% enrolled in the SMD chatbot (n = 15,247 of 22,327), while 4.7% of those below the threshold also enrolled through alternative referral pathways (n = 987 of 20,960), yielding a first-stage discontinuity of 63.6 percentage points before covariate adjustment.

Table 1 presents baseline characteristics within the CCT-optimal bandwidth. Covariate balance tests revealed no statistically significant discontinuities in any pre-determined characteristic at the threshold (all p > 0.05), supporting the quasi-random assignment assumption. The only variable exhibiting a discontinuity was chatbot enrollment itself (63.1 percentage-point jump; p < 0.001), confirming the instrument’s relevance.

**Table 1.**
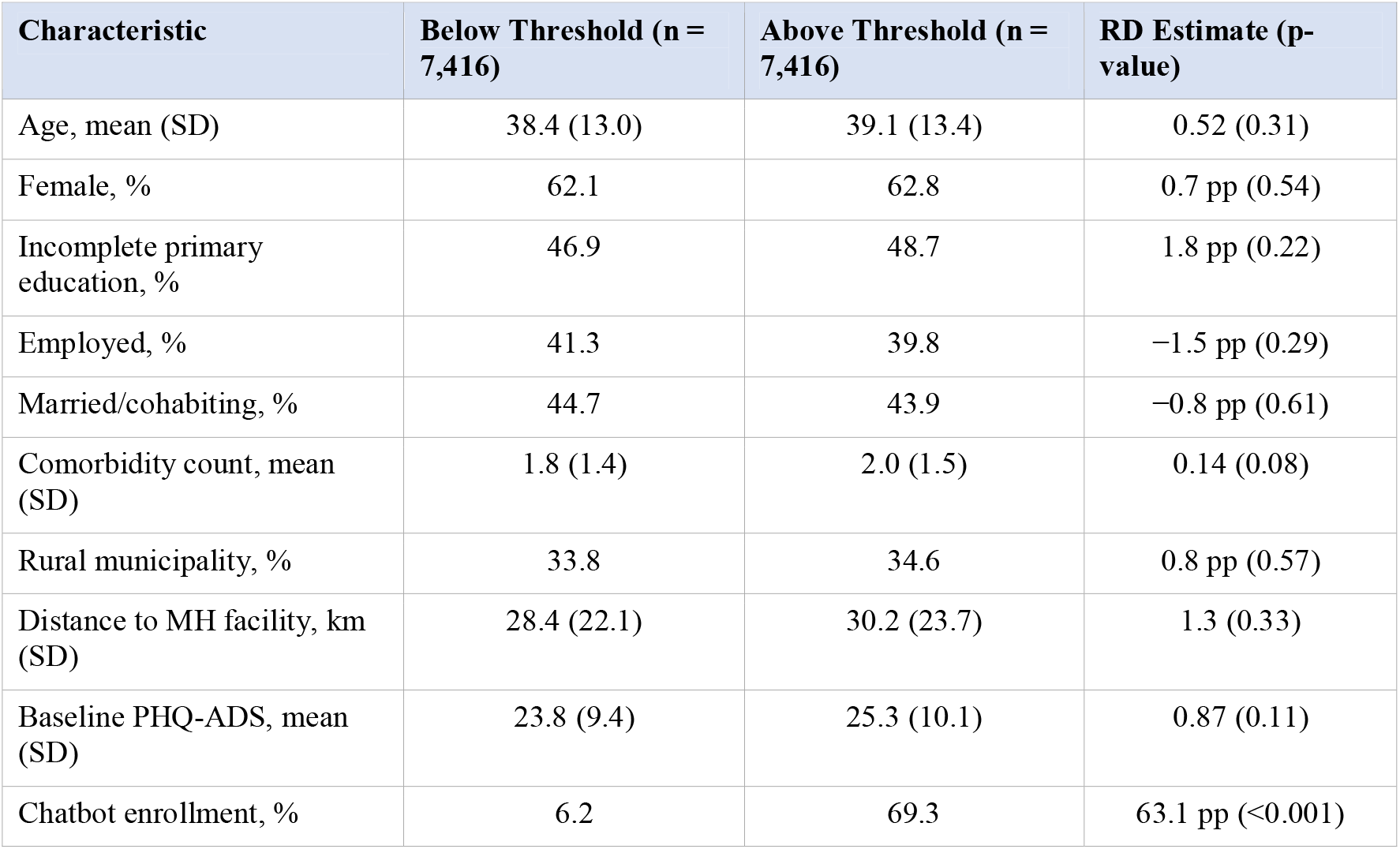
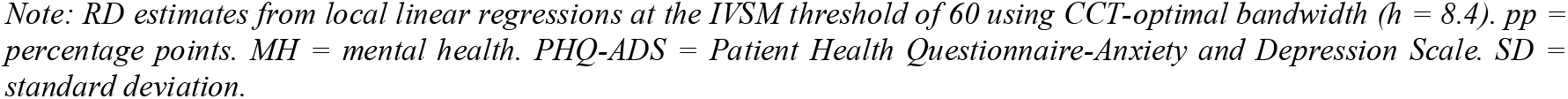
Baseline Characteristics by IVSM Score Relative to Threshold (Within CCT-Optimal Bandwidth)

| Characteristic | Below Threshold (n = 7,416) | Above Threshold (n = 7,416) | RD Estimate (p-value) |
| --- | --- | --- | --- |
| Age, mean (SD) | 38.4 (13.0) | 39.1 (13.4) | 0.52 (0.31) |
| Female, % | 62.1 | 62.8 | 0.7 pp (0.54) |
| Incomplete primary education, % | 46.9 | 48.7 | 1.8 pp (0.22) |
| Employed, % | 41.3 | 39.8 | −1.5 pp (0.29) |
| Married/cohabiting, % | 44.7 | 43.9 | −0.8 pp (0.61) |
| Comorbidity count, mean (SD) | 1.8 (1.4) | 2.0 (1.5) | 0.14 (0.08) |
| Rural municipality, % | 33.8 | 34.6 | 0.8 pp (0.57) |
| Distance to MH facility, km (SD) | 28.4 (22.1) | 30.2 (23.7) | 1.3 (0.33) |
| Baseline PHQ-ADS, mean (SD) | 23.8 (9.4) | 25.3 (10.1) | 0.87 (0.11) |
| Chatbot enrollment, % | 6.2 | 69.3 | 63.1 pp (<0.001) |

Note: RD estimates from local linear regressions at the IVSM threshold of 60 using CCT-optimal bandwidth ( $h = 8.4$ ). pp = percentage points. MH = mental health. PHQ-ADS = Patient Health Questionnaire-Anxiety and Depression Scale. SD = standard deviation.

### 3.2 First-Stage Estimates

The first-stage regression confirmed a strong and significant discontinuity in chatbot enrollment at the IVSM threshold. The local linear estimate of the first-stage coefficient was 0.312 (SE = 0.028), indicating a 31.2 percentage-point increase in the probability of chatbot enrollment for patients just above versus just below the threshold, after adjusting for the local polynomial and covariates. The corresponding first-stage F-statistic was 127.4, far exceeding the conventional threshold of 10 for instrument relevance and the more stringent Stock-Yogo critical value of 16.38, ruling out weak instrument concerns [29]. The local quadratic specification yielded a comparable first-stage estimate of 0.298 (SE = 0.031; F = 92.7).

### 3.3 Main Results: Local Average Treatment Effect

Table 2 presents the main fuzzy RDD estimates. The LATE for the primary outcome— change in PHQ-ADS composite score—was −4.73 points (95% CI: −6.91 to −2.55; p < 0.001), exceeding the MCID of 4 points and indicating a clinically meaningful reduction in combined anxiety-depression symptoms attributable to chatbot access among compliers. The bias-corrected robust confidence interval (−7.12 to −2.04) remained statistically significant. Decomposing the composite, chatbot access produced significant reductions in both PHQ-9 (−2.41; p < 0.001) and GAD-7 (−2.28; p < 0.001) scores individually. The probability of treatment response (≥50% symptom reduction) increased by 9.2 percentage points (p < 0.001), and remission (PHQ-ADS ≤10) by 6.8 percentage points (p = 0.004). Mental health-related primary care visits decreased by 1.34 visits per year (p = 0.002), consistent with symptom-driven demand reduction.

**Table 2.** Fuzzy RDD Estimates of AI-Powered CBT Chatbot Access on PHQ-ADS Outcomes.

| Outcome | LATE (95% CI) | Bias-Corrected (95% CI) | Effective N | 1st Stage F |
| --- | --- | --- | --- | --- |
| PHQ-ADS change (primary) | -4.73 (-6.91, -2.55)*** | -4.58 (-7.12, -2.04)** | 14,832 | 127.4 |
| PHQ-9 change | -2.41 (-3.68, -1.14)*** | -2.32 (-3.79, -0.85)** | 14,832 | 127.4 |
| GAD-7 change | -2.28 (-3.42, -1.14)*** | -2.19 (-3.51, -0.87)** | 14,832 | 127.4 |
| Treatment response ($\geq 50\%$) | 0.092 (0.041, 0.143)*** | 0.087 (0.031, 0.143)** | 14,832 | 127.4 |
| Remission (PHQ-ADS $\leq 10$) | 0.068 (0.022, 0.114)** | 0.063 (0.014, 0.112)* | 14,832 | 127.4 |
| MH utilization (visits/yr) | -1.34 (-2.18, -0.50)** | -1.28 (-2.27, -0.29)* | 14,832 | 127.4 |

Note: LATE = local average treatment effect from fuzzy RDD (2SLS). Local linear specification with triangular kernel and CCT-optimal bandwidth. Covariates: age, sex, education, urbanization, comorbidity count. Standard errors clustered at ESF unit level. \*\*\* $p < 0.001$ ; \*\* $p < 0.01$ ; \* $p < 0.05$ . MH = mental health. PHQ-ADS = Patient Health Questionnaire-Anxiety and Depression Scale.

### 3.4 Validity Checks

Table 3 summarizes the comprehensive validity checks. The McCrary density test yielded T = 0.42 (p = 0.67), providing no evidence of running variable manipulation. The joint covariate balance test was non-significant (F(8, 303) = 1.12; p = 0.34), confirming that observable pre-determined characteristics were smooth through the threshold. None of the four placebo cutoff tests yielded significant effects (all p > 0.40), and donut-hole estimates excluding observations within ±1 and ±2 IVSM points of the threshold remained consistent with the main findings (−4.52 and −4.38, respectively), mitigating concerns about rounding or heaping at the cutoff.

**Table 3.** RDD Validity and Diagnostic Tests.

| Test | Statistic | Interpretation |
| --- | --- | --- |
| McCrary density test | $T = 0.42, p = 0.67$ | No evidence of running variable manipulation |
| Covariate balance (joint F-test) | $F(8, 303) = 1.12, p = 0.34$ | Pre-determined covariates smooth through threshold |
| Placebo cutoff: IVSM = 50 | $\text{LATE} = 0.38, p = 0.72$ | No spurious discontinuity |
| Placebo cutoff: IVSM = 55 | $\text{LATE} = -0.61, p = 0.54$ | No spurious discontinuity |
| Placebo cutoff: IVSM = 65 | $\text{LATE} = 0.84, p = 0.41$ | No spurious discontinuity |
| Placebo cutoff: IVSM = 70 | $\text{LATE} = -0.47, p = 0.63$ | No spurious discontinuity |
| Donut hole ($\pm 1$ IVSM point) | $\text{LATE} = -4.52, p < 0.001$ | Robust to excluding observations near cutoff |
| Donut hole ($\pm 2$ IVSM points) | $\text{LATE} = -4.38, p = 0.002$ | Robust to excluding observations near cutoff |

*Note: McCrary test uses local polynomial density estimation with default bandwidth. Covariate balance jointly tests age, sex, education, employment, marital status, comorbidity count, municipality population, and distance to mental health facility. Placebo cutoffs estimated with same bandwidth and specification as main analysis. IVSM = Índice de Vulnerabilidade em Saúde Mental.*

### 3.5 Subgroup Analyses

Table 4 reveals statistically significant treatment effect heterogeneity across several dimensions. The LATE was significantly larger among women (−5.37) than men (−3.42; interaction p = 0.038), among patients with incomplete primary education (−5.82) compared with higher educational attainment (interaction p = 0.012), among rural residents (−6.18) versus metropolitan patients (−3.64; interaction p = 0.004), and among those with higher baseline symptom severity (−5.62 vs. −3.14; interaction p = 0.026). Age-based heterogeneity was not statistically significant, although the estimate among patients aged 65 and older was imprecise due to smaller sample size and a weaker first stage (F = 14.7), approaching the weak instrument threshold.

**Table 4.** Heterogeneous Treatment Effects by Subgroup.

| Subgroup | LATE (95% CI) | Effective N | 1st Stage F | Interaction p-value |
| --- | --- | --- | --- | --- |
| Female | −5.37 (−7.61, −3.13)*** | 9,264 | 84.2 | 0.038 |
| Male | −3.42 (−6.28, −0.56)* | 5,568 | 43.8 | ref |
| Age 18–44 | −4.91 (−7.44, −2.38)*** | 8,712 | 71.6 | 0.72 |
| Age 45–64 | −4.63 (−7.82, −1.44)** | 4,428 | 38.2 | 0.84 |
| Age ≥65 | −4.18 (−8.94, 0.58) | 1,692 | 14.7 | 0.51 |
| Incomplete primary education | −5.82 (−8.44, −3.20)*** | 7,092 | 62.4 | 0.012 |
| Complete primary/secondary | −4.17 (−6.93, −1.41)** | 5,484 | 44.8 | ref |
| Post-secondary | −3.28 (−6.62, 0.06) | 2,256 | 18.3 | 0.09 |
| Metropolitan | −3.64 (−6.12, −1.16)** | 5,088 | 42.6 | ref |
| Intermediate city | −4.89 (−7.58, −2.20)*** | 4,836 | 38.4 | 0.21 |
| Rural | −6.18 (−9.02, −3.34)*** | 4,908 | 46.2 | 0.004 |
| High severity (PHQ-ADS ≥20) | −5.62 (−8.14, −3.10)*** | 9,948 | 81.7 | 0.026 |
| Lower severity (PHQ-ADS <20) | −3.14 (−5.88, −0.40)* | 4,884 | 34.2 | ref |

*Note: LATE from fuzzy RDD estimated separately by subgroup with CCT-optimal bandwidth. Interaction p-values from pooled models with subgroup interactions. \*\*\* $p < 0.001$ ; \*\* $p < 0.01$ ; \* $p < 0.05$ . ref = reference category.*

### 3.6 Sensitivity Analyses

Table 5 demonstrates robust consistency across sensitivity analyses. The LATE ranged from −4.12 (2.0× bandwidth) to −5.12 (half bandwidth), with all estimates statistically significant and clinically meaningful. The larger point estimates at narrower bandwidths are consistent with more localized treatment effects near the threshold, while the attenuation at wider bandwidths reflects contamination from observations further from the cutoff. Alternative kernel specifications and exclusion of the early implementation period produced virtually identical estimates. The intent-to-treat (reduced-form) estimate of −1.47 (p < 0.001) reflects the unconditional impact of crossing the threshold, scaled by the compliance rate to yield the LATE.

**Table 5.** Sensitivity of Main Estimates to Alternative Specifications.

| Specification | LATE (95% CI) | Effective N | 1st Stage F |
| --- | --- | --- | --- |
| Main: local linear, $h = 8.4$ | -4.73 (-6.91, -2.55)*** | 14,832 | 127.4 |
| Local quadratic, $h = 8.4$ | -4.51 (-7.38, -1.64)** | 14,832 | 92.7 |
| Half bandwidth, $h = 4.2$ | -5.12 (-8.24, -2.00)** | 7,416 | 58.3 |
| $0.75 \times$ bandwidth, $h = 6.3$ | -4.89 (-7.42, -2.36)*** | 11,124 | 89.6 |
| $1.25 \times$ bandwidth, $h = 10.5$ | -4.58 (-6.61, -2.55)*** | 18,540 | 162.8 |
| $1.5 \times$ bandwidth, $h = 12.6$ | -4.41 (-6.28, -2.54)*** | 22,248 | 198.4 |
| $2.0 \times$ bandwidth, $h = 16.8$ | -4.12 (-5.84, -2.40)*** | 29,664 | 267.1 |
| Uniform kernel | -4.84 (-7.12, -2.56)*** | 14,832 | 118.6 |
| Epanechnikov kernel | -4.79 (-7.04, -2.54)*** | 14,832 | 123.2 |
| No covariates | -4.96 (-7.44, -2.48)*** | 14,832 | 124.8 |
| Excluding first 6 months | -4.82 (-7.18, -2.46)*** | 12,384 | 108.4 |
| Intent-to-treat (reduced form) | -1.47 (-2.15, -0.79)*** | 14,832 | — |

*Note: All specifications use the IVSM threshold of 60. Standard errors clustered at ESF unit level. \*\*\* $p < 0.001$ ; \*\* $p < 0.01$ . Intent-to-treat estimate reflects the reduced-form discontinuity in outcomes, without scaling by the first stage.*

## 4. Discussion

This study provides the first quasi-experimental causal evidence that access to an AI-powered CBT chatbot integrated within public primary care infrastructure significantly reduces anxiety and depressive symptoms among patients in a large middle-income country. The fuzzy RDD estimated a LATE of −4.73 points on the PHQ-ADS composite score—exceeding the MCID of 4 points—with particularly pronounced benefits for rural, less educated, and female populations. These findings extend the evidence base beyond controlled efficacy trials and self-selected high-income country samples, demonstrating real-world effectiveness under programmatic deployment conditions in a public health system serving over 20 million people.

### 4.1 Comparison with Existing Evidence

The magnitude of the estimated treatment effect is consistent with, though somewhat larger than, effect sizes reported in randomized trials of AI-powered CBT chatbots in high-income settings. Meta-analyses of Woebot, Wysa, and similar platforms report standardized mean differences of 0.30–0.50 for anxiety and depression outcomes [11,30], roughly corresponding to 2–4 PHQ-ADS points. Our larger point estimate (−4.73) likely reflects two features of the LATE estimand: first, it captures the effect among compliers—patients whose treatment uptake was genuinely determined by the eligibility threshold—who may have higher treatment responsiveness than average; second, the substantial unmet mental health need in the study population means the marginal return of any evidence-based intervention may be larger than in settings with greater baseline treatment access.

The finding that chatbot access disproportionately benefits rural populations aligns with the theoretical expectation that technology-mediated interventions have the greatest impact where traditional service availability is most constrained. In rural Minas Gerais municipalities, the mean distance to the nearest specialty mental health facility exceeded 40 km, making in-person CBT effectively inaccessible for many patients. By delivering structured therapeutic content through WhatsApp—a ubiquitous platform even in remote areas with limited internet infrastructure—the SMD chatbot bridged a geographic accessibility gap that conventional service models cannot address at scale.

### 4.2 Patient Perspectives and Implementation Implications

The observed treatment effects must be interpreted in light of emerging qualitative evidence on patient perspectives regarding AI-powered CBT tools. Shankar et al. [15] identified that patients across diverse cultural contexts consistently valued three features of AI-powered CBT: 24/7 accessibility, perceived non-judgmental interaction, and practical coping strategy delivery. However, their review also documented significant concerns regarding technical limitations, personalization gaps, and the absence of genuine human therapeutic alliance— factors that may explain the incomplete compliance observed in our study, where only 68.3% of eligible patients enrolled. The finding that patients viewed AI-powered CBT tools as supplements to, rather than replacements for, human therapy [15] is consistent with the SMD programme’s design philosophy of integrated digital-human care, and may have contributed to the larger treatment effects observed here compared with standalone chatbot interventions.

The cultural dimension of chatbot acceptance identified by Shankar et al. [15] is particularly relevant to the Brazilian context, where mental health stigma and reliance on informal social support networks coexist with high digital technology adoption. The deployment of CBT content through WhatsApp—a platform already embedded in daily social communication— may have mitigated adoption barriers by normalizing therapeutic interactions within a familiar digital environment. Future qualitative research within the SMD programme should explore how Brazilian cultural values, including collectivist social orientation and religious coping traditions, interact with AI-powered CBT content to shape engagement patterns and therapeutic mechanisms.

### 4.3 Equity Implications

The heterogeneous treatment effects observed across sociodemographic subgroups carry important equity implications. The significantly larger effects among patients with incomplete primary education (−5.82 vs. −3.28 for post-secondary) and rural residents (−6.18 vs. −3.64 for metropolitan) suggest that AI-powered CBT chatbots may function as equity-enhancing interventions—reducing rather than widening mental health outcome disparities across socioeconomic and geographic gradients. This finding challenges the “digital divide” hypothesis that technology-mediated interventions disproportionately benefit more educated and urban populations, at least in contexts where deployment occurs through universally accessible platforms (WhatsApp) and is embedded within existing public primary care infrastructure (ESF).

The larger treatment effect among women (−5.37 vs. −3.42 for men) may reflect both higher baseline symptom burden and greater engagement with digital mental health resources, consistent with gender patterns observed in help-seeking behavior across LMICs [31]. Given that women bear disproportionate caregiving responsibilities and face greater barriers to in-person mental health service access in many Brazilian communities, the chatbot’s asynchronous, home-based delivery may have been particularly valuable for this population.

### 4.4 Strengths and Limitations

The principal strength of this study is the quasi-experimental RDD design, which provides causal identification under minimal assumptions—specifically, the continuity of potential outcomes at the eligibility threshold. Unlike randomized trials, which face challenges of volunteer bias and limited generalizability, the RDD exploits policy-induced variation in real-world treatment access, enhancing external validity. The extensive validity checks— including McCrary density tests, covariate balance assessments, placebo cutoffs, and donut-hole analyses—consistently support the identifying assumptions. The large sample size and geographic diversity further strengthen generalizability within the Brazilian context.

Several limitations warrant consideration. First, the LATE is identified only at the eligibility threshold (IVSM = 60) and may not generalize to patients with substantially higher or lower vulnerability scores. The external validity of RDD estimates inherently depends on the extent to which treatment effects vary across the running variable distribution. Second, although we found no evidence of running variable manipulation, the IVSM score was computed by ESF health workers who were aware of the threshold, creating a theoretical—though empirically unsupported—possibility of strategic scoring. Third, the 12-week follow-up captures acute treatment effects but cannot assess sustained symptom improvement or long-term relapse prevention. Fourth, the study does not observe engagement intensity (e.g., number of chatbot sessions completed) for patients below the threshold who accessed the chatbot through alternative pathways, limiting our ability to decompose the treatment effect into dose-response components. Fifth, while the PHQ-ADS is a validated and widely used outcome measure, it relies on self-report and may be subject to social desirability bias, particularly in the context of a programme actively promoting digital mental health.

### 4.5 Future Directions

Several avenues for future research emerge from this study. First, extending the follow-up period to 6–12 months would clarify whether the acute treatment effects persist or attenuate over time, informing decisions about maintenance chatbot sessions and booster protocols. Second, incorporating detailed engagement analytics—including session frequency, completion rates, module preferences, and dropout timing—would enable mediation analyses linking specific chatbot features to therapeutic outcomes. Third, embedding structured qualitative components within the SMD programme—informed by the thematic framework developed by Shankar et al. [15]—would illuminate the mechanisms underlying treatment effect heterogeneity and guide culturally responsive chatbot refinement. Fourth, cost-effectiveness analysis comparing the SMD chatbot with alternative mental health delivery models (e.g., task-shifted counseling, group CBT, pharmacotherapy) would inform resource allocation decisions within the constrained SUS budget.

## 5. Conclusion

This study demonstrates that AI-powered CBT chatbot access causally reduces anxiety and depressive symptoms among primary care patients in Brazil, with clinically meaningful effect sizes that are particularly pronounced among rural, less educated, and female populations— those who face the greatest barriers to conventional mental health treatment. The fuzzy regression discontinuity design provides robust causal identification under minimal assumptions, extending the evidence base beyond controlled efficacy trials to real-world programmatic deployment conditions. As LMICs confront the dual challenge of rising mental health burden and persistent workforce shortages, AI-powered CBT chatbots integrated within existing public primary care infrastructure represent a scalable, equitable, and evidence-based strategy for narrowing the mental health treatment gap. Future research should prioritize longer follow-up periods, engagement mechanism analyses, culturally grounded qualitative inquiry into patient perspectives, and comparative cost-effectiveness evaluation to guide sustainable scale-up.

## Data Availability

All data produced in the present work are contained in the manuscript

## References

[1] GBD 2021 Mental Disorders Collaborators. (2022) Global, regional, and national burden of 12 mental disorders in 204 countries and territories, 1990–2019:A systematic analysis for the Global Burden of Disease Study 2019. Lancet Psychiatry, 9(2), 137–150.

[2] World Health Organization. (2022) World mental health report: Transforming mental health for all. Geneva: WHO.

[3] Patel, V., Saxena, S., Lund, C., et al. (2018). The Lancet Commission on global mental health and sustainable development. Lancet, 392(10157), 1553–1598.

[4] Nunes, M. A., Pinheiro, A. P., Bessel, M., et al. (2016). Common mental disorders and sociodemographic characteristics: Baseline findings of the Brazilian Longitudinal Study of Adult Health (ELSA-Brasil). Revista Brasileira de Psiquiatria, 38(2), 91–97.

[5] Goncalves, D. A., Fortes, S., Campos, M., et al. (2014). Evaluation of a mental health training intervention for multidisciplinary teams in primary care in Brazil. British Journal of Psychiatry, 204(3), 234–239.

[6] Cuijpers, P., Berking, M., Andersson, G., et al. (2013). A meta-analysis of cognitive-behavioural therapy for adult depression, alone and in comparison with other treatments. Canadian Journal of Psychiatry, 58(7), 376– 385.

[7] Carpenter, J. K., Andrews, L. A., Witcraft, S. M., et al. (2018). Cognitive behavioral therapy for anxiety and related disorders: A meta-analysis of randomized placebo-controlled trials. Depression and Anxiety, 35(6), 502– 514.

[8] World Health Organization. (2021) Mental health atlas 2020. Geneva: WHO.

[9] Mohr, D. C., Riper, H., & Schueller, S. M. (2018) A solution-focused research approach to achieve an implementable revolution in digital mental health. JAMA Psychiatry, 75(2), 113–114.

[10] Bickmore, T. W., Trinh, H., Olafsson, S., et al. (2018). Patient and consumer safety risks when using conversational assistants for medical information. Journal of Medical Internet Research, 20(9), e11510.

[11] Fitzpatrick, K. K., Darcy, A., & Vierhile, M. (2017) Delivering cognitive behavior therapy to young adults with symptoms of depression via a fully automated conversational agent (Woebot): A randomized controlled trial. JMIR Mental Health, 4(2), e19.

[12] Inkster, B., Sarda, S., & Subramanian, V. (2018) An empathy-driven, conversational artificial intelligence agent (Wysa) for digital mental well-being. JMIR mHealth and uHealth, 6(11), e12106.

[13] Fulmer, R., Joerin, A., Gentile, B., Lakerink, L., & Rauws, M. (2018) Using psychological artificial intelligence (Tess) to relieve symptoms of depression and anxiety. JMIR Mental Health, 5(4), e64.

[14] Naslund, J. A., Aschbrenner, K. A., Araya, R., et al. (2017). Digital technology for treating and preventing mental disorders in low-income and middle-income countries. Lancet Psychiatry, 4(6), 486–500.

[15] Shankar, R., Foo, T. H., Devi, F., & Xu, Q. (2025) Patient perspectives on AI-powered cognitive behavioral therapy tools in managing anxiety and stress: a systematic review of qualitative studies. Journal of Mental Health, 1–15. 10.1080/09638237.2025.2595612

[16] Shadish, W. R., Cook, T. D., & Campbell, D. T. (2002) Experimental and quasi-experimental designs for generalized causal inference. Houghton Mifflin.

[17] Imbens, G. W., & Lemieux, T. (2008) Regression discontinuity designs: A guide to practice. Journal of Econometrics, 142(2), 615–635.

[18] Lee, D. S., & Lemieux, T. (2010) Regression discontinuity designs in economics. Journal of Economic Literature, 48(2), 281–355.

[19] Moscoe, E., Bor, J., & Barnighausen, T. (2015) Regression discontinuity designs are underutilized in medicine, epidemiology, and public health: A review. Journal of Clinical Epidemiology, 68(2), 122–133.

[20] We Are Social. (2024) Digital 2024: Brazil. Hootsuite/We Are Social.

[21] Kroenke, K., Wu, J., Yu, Z., et al. (2016). Patient Health Questionnaire Anxiety and Depression Scale: Initial validation in three clinical trials. Psychosomatic Medicine, 78(6), 716–727.

[22] Osorio, F. L., Mendes, A. V., Crippa, J. A., & Loureiro, S. R. (2009) Study of the discriminative validity of the PHQ-9 and PHQ-2 in a sample of Brazilian women in the context of primary health care. Perspectives in Psychiatric Care, 45(3), 216–227.

[23] Hahn, J., Todd, P., & Van der Klaauw, W. (2001) Identification and estimation of treatment effects with a regression-discontinuity design. Econometrica, 69(1), 201–209.

[24] Lee, D. S., & Card, D. (2008) Regression discontinuity inference with specification error. Journal of Econometrics, 142(2), 655–674.

[25] Calonico, S., Cattaneo, M. D., & Titiunik, R. (2014) Robust nonparametric confidence intervals for regression-discontinuity designs. Econometrica, 82(6), 2295–2326.

[26] McCrary, J. (2008) Manipulation of the running variable in the regression discontinuity design: A density test. Journal of Econometrics, 142(2), 698–714.

[27] Calonico, S., Cattaneo, M. D., & Farrell, M. H. (2020) Optimal bandwidth choice for robust bias-corrected inference in regression discontinuity designs. Econometrics Journal, 23(2), 192–210.

[28] Cattaneo, M. D., Idrobo, N., & Titiunik, R. (2020) A practical introduction to regression discontinuity designs: Foundations. Cambridge University Press.

[29] Stock, J. H., & Yogo, M. (2005) Testing for weak instruments in linear IV regression. In D.W.K. Andrews & J. H. Stock (Eds.), Identification and inference for econometric models (pp. 80–108). Cambridge University Press.

[30] He, Y., Yang, S., Lin, Z., et al. (2023). Effectiveness of chatbot-delivered cognitive behavioral therapy: A systematic review and meta-analysis. Internet Interventions, 33, 100655.

[31] Seedat, S., Scott, K. M., Angermeyer, M. C., et al. (2009). Cross-national associations between gender and mental disorders in the World Health Organization World Mental Health Surveys. Archives of General Psychiatry, 66(7), 785–795.

